# Shared respiratory infectious disease hotspots identify priority countries for pandemic preparedness: a Bayesian spatiotemporal analysis with COVID-19 external validation

**DOI:** 10.64898/2026.07.09.26357662

**Authors:** Qiancheng Ma, Tianzhen Zhang, Dongzhi Lin

## Abstract

To identify countries with potential weaknesses in respiratory public health protection, we characterised shared hotspot patterns across three major respiratory infectious diseases and assessed whether the resulting shared hotspot scores were associated with worse COVID-19 outcomes. A Bayesian multivariate shared-component spatiotemporal model was fitted to data from 204 countries over 1990–2023 using Global Burden of Disease 2023 estimates to derive a shared hotspot score for each country. Generalized estimating equation negative binomial models were then used to examine associations between the shared hotspot score and COVID-19 incidence and mortality over 2020–2023. The shared hotspot score showed substantial cross-country heterogeneity, with the highest values concentrated in sub-Saharan Africa, South Asia, and Southeast Asia. Tuberculosis showed the strongest contribution to the shared spatial component (λ = 1.657, 95% highest density interval: 0.883–2.506). Higher shared hotspot scores were significantly associated with both higher COVID-19 incidence (incidence rate ratio = 1.6783, 95% confidence interval: 1.4564–1.9340; p = 8.308 × 10□13) and mortality (incidence rate ratio = 1.7436, 95% confidence interval: 1.5061–2.0186; p = 9.912 × 10□14). Countries with persistently high co-occurrence of common respiratory infectious diseases also experienced worse COVID-19 outcomes, suggesting that the shared hotspot score may inform preparedness-oriented surveillance and resource allocation for future large-scale respiratory epidemics or pandemics.

## 1. Introduction

COVID-19 caused substantial mortality, profound social disruption, and major economic losses worldwide. Although the global emergency phase was declared over in 2023, the pandemic highlighted how unevenly countries were able to prevent, absorb, and respond to major respiratory epidemics. This unevenness matters because future large-scale respiratory epidemics are unlikely to be avoided entirely, and global control may be delayed when a subset of countries remains persistently vulnerable. Identifying such countries before the next major respiratory pandemic may therefore be critical for prioritising support and improving the efficiency of global preparedness efforts.

Upper respiratory infections (URIs) involve the upper airways and are commonly caused by respiratory viruses such as rhinoviruses, common human coronaviruses, adenoviruses, parainfluenza viruses, enteroviruses and human metapneumovirus[1]. Lower respiratory infections (LRIs) involve the lower respiratory tract, from the trachea and bronchi to the bronchioles and alveoli, and are commonly caused by pathogens such as Streptococcus pneumoniae, Haemophilus influenzae, respiratory syncytial virus (RSV), and influenza viruses[2, 3]. Tuberculosis (TB) is an airborne infectious disease caused by Mycobacterium tuberculosis that most often affects the lungs[4]. Because pulmonary TB is transmitted through the air and remains a major cause of respiratory infectious morbidity and mortality worldwide [4, 5], the control patterns of these three diseases may reflect a country’s underlying capacity to prevent and manage respiratory infectious diseases.

Previous work on respiratory pandemic preparedness has mainly focused on preparedness frameworks, simulation exercises, or written national plans[6–8]. Although these studies provide useful information on functional capacities and planning gaps, they do not directly assess whether countries show persistent vulnerability across multiple common respiratory infectious diseases in real-world settings. They also rarely examine whether such vulnerability signals are supported by external pandemic outcomes.

In this study, a Bayesian shared hotspot model was first used to identify the common geographic hotspot patterns of three common respiratory infectious diseases across countries. Specifically, we modelled upper respiratory infections, lower respiratory infections, and tuberculosis jointly, and derived a shared hotspot score to quantify the extent to which each country was involved in a common respiratory infectious disease hotspot pattern. This study then conducted an external validation analysis using country-level COVID-19 data to examine whether countries with higher shared hotspot scores performed worse during the pandemic, as reflected by higher COVID-19 incidence and mortality.

Compared with preparedness studies based mainly on frameworks, simulation exercises, or COVID-19 alone, this study uses long-term observed data (1990–2023), jointly analyses three common respiratory infectious diseases, and externally validates the shared hotspot score using real-world COVID-19 outcomes.

## 2. Materials and methods

### 2.1. Software and data processing

All data processing and statistical analyses were performed using Python (version 3.13.9) within the Anaconda environment. Figures were generated and spatial visualisation was conducted in Python using GeoPandas and Matplotlib.

### 2.2. Data sources

Annual country-level incidence data for tuberculosis, lower respiratory infections, and upper respiratory infections were obtained from the Global Burden of Disease (GBD) 2023 study[9]. Country-year population data were obtained from the World Bank[10]. All 204 countries with harmonized data across the required sources were included in the analysis. Country-level socioeconomic, health-system, and environmental indicators, including gross domestic product per capita, population density, current health expenditure per capita, ambient PM _2.5_ concentration, physician density, and the Universal Health Coverage (UHC) service coverage index, were compiled from World Bank sources[11–16]. July temperature and July humidity were obtained from the World Bank Climate Change Knowledge Portal[17].

### 2.3. Study design and analytical framework

A Bayesian multivariate shared-component spatiotemporal negative binomial model was fitted to jointly analyse the three diseases. The shared-component framework was adapted to annual country-level count data for three diseases, with a negative binomial likelihood, scaled ICAR spatial structure, disease-specific RW(1) temporal effects, and standardized country-level covariates[18–24]. The main model was specified as:

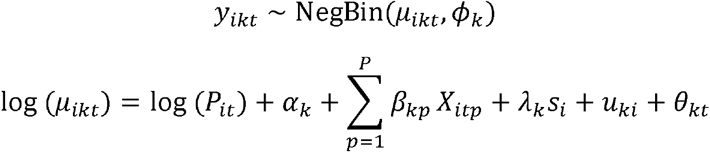

Where:*Y*_*ikt*_ is the observed count for country *i*, disease *k*, and year *t*; *μ*_*ikt*_ is the expected count; *⌽*_*k*_ is the disease-specific overdispersion parameter of the negative binomial distribution. *P*_*it*_ is the population offset; *α*_*k*_ is the disease-specific intercept; *X*_*itp*_ is the *p*-th covariate and *β*_*kp*_ its disease-specific coefficient; *s*_*i*_ is the shared spatial component; *λ*_*k*_ is the disease-specific loading; *u*_*ki*_ is the disease-specific spatial deviation; and *θ*_*kt*_ is the disease-specific temporal effect, modeled as a first-order random walk.

For external validation, generalized estimating equation negative binomial models were fitted for annual COVID-19 incidence and mortality[21, 25]. The external validation model was specified as:

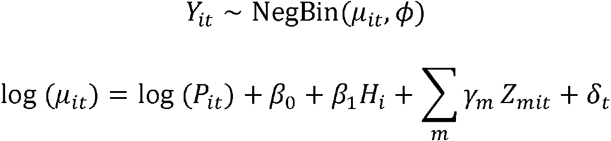

Where: *Y*_*it*_ denotes the annual COVID-19 outcome count (incidence or deaths) for country *i* in year *t*, modeled separately for incident cases and deaths; *μ*_*it*_ is the expected count; log (*P*_*it*_)is the POPULATION offset; *H*_*i*_ is the standardized shared hotspot score; *Z*_*mit*_ denotes the *m*-th time-varying covariate; *γ*_*m*_is its corresponding coefficient; *δ*_*t*_ represents year fixed effects; and *β*_1_ is the coefficient of primary interest.

### 2.4. Derivation of the shared hotspot score

In this model, the shared spatial component *s*_*i*_ represents a time-invariant latent geographic structure common to all three diseases, while temporal variation is captured separately by the disease-specific random walk *θ*_*kt*_. Accordingly, *s*_*i*_ reflects the long-term average shared spatial pattern across the full study period (1990–2023), rather than a snapshot from any single year.

The country-level shared hotspot score *H*_*i*_ was derived from the posterior distributions of *s*_*i*_ and *λ*_*k*_ as a loading-weighted summary:

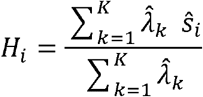

where 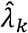 and *ŝ*_*i*_ denote the posterior means of the disease-specific loading and the shared spatial component, respectively. Because *s*_*i*_ does not vary across diseases, this expression reduces to *H*_*i*_ = *ŝ*_*i*_. Positive values indicate countries whose respiratory disease geography is consistent with a shared hotspot pattern; negative values indicate departure from it. Before entering the external validation model, *H*_*i*_ was standardized to zero mean and unit variance so that the resulting incidence rate ratios reflect a one-standard-deviation increase.

A schematic overview of the analytical process is shown in Fig 1, and the detailed model specifications are provided in Supporting information S1 Text.

**Fig 1.**
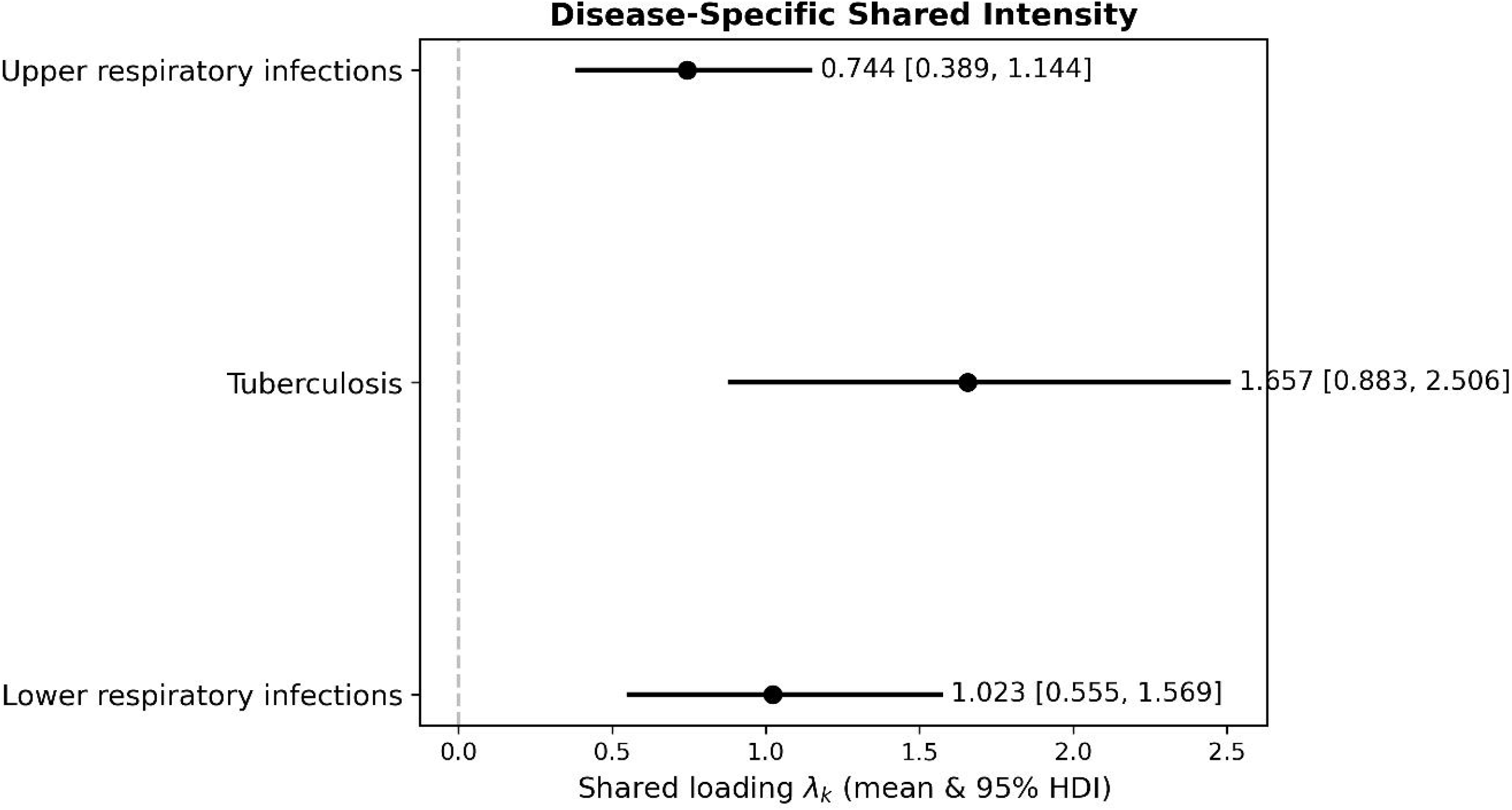
Analytical framework of the study

All data processing and statistical analyses were conducted in Python 3.13.9 within the Anaconda environment, and spatial data processing and mapping were performed in ArcGIS Pro 3.5.2.

## 3. Results

### 3.1. Descriptive characteristics of the study panel

#### 3.1.1. Descriptive characteristics of the three-disease study panel

The three-disease study panel included 204 countries, with annual observations spanning 1990 to 2023, yielding 6,936 country-year observations for each disease series. Across the full study period, the cumulative number of cases was 343,945,799 for tuberculosis, 8,382,572,859 for lower respiratory infections, and 390,446,114,133 for upper respiratory infections. Among the three diseases, upper respiratory infections contributed the largest cumulative burden, followed by lower respiratory infections, whereas tuberculosis had the smallest total burden. The annual aggregate trends of these three diseases are shown in Fig 2.

**Fig 2.**
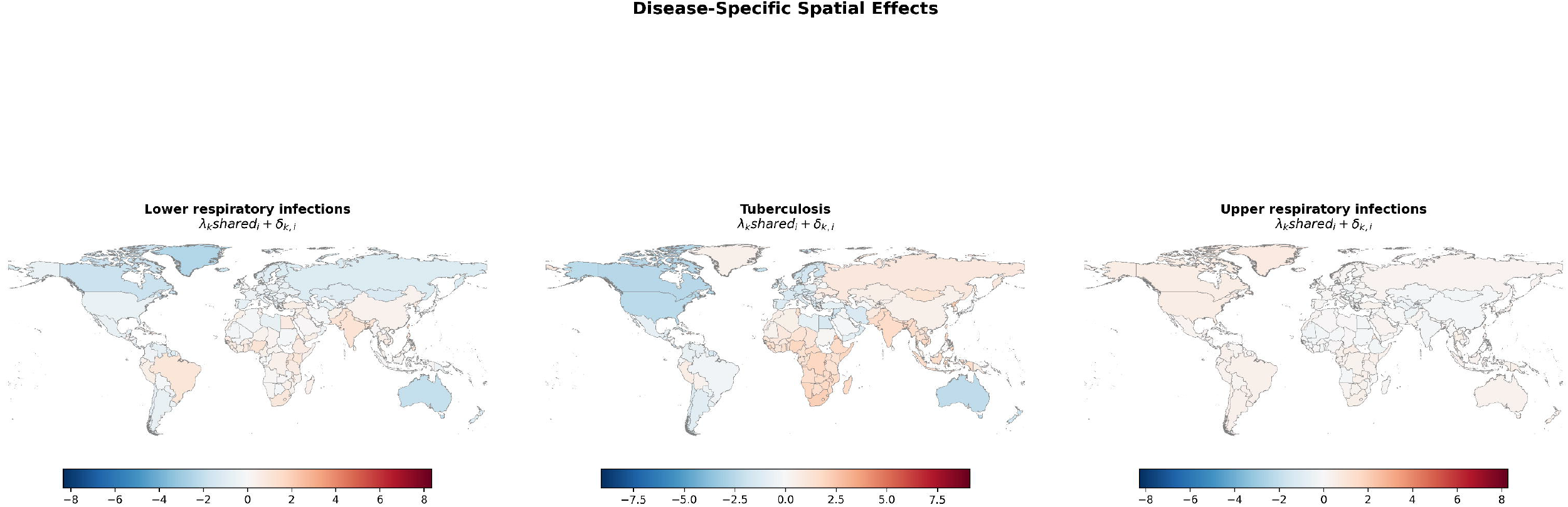
Annual global aggregated case counts for three common respiratory infectious diseases and COVID-19 outcomes, 1990–2023

#### 3.1.2. Descriptive characteristics of the COVID-19 validation panel

The harmonized country-year dataset for external validation covered 204 countries from 2020 to 2023, yielding 816 country-year observations for each COVID-19 outcome series. The annual trajectories of total COVID-19 cases and deaths are presented in Fig 2.

### 3.2. Diagnostics and robustness of the shared hotspot model

To evaluate the adequacy and robustness of the shared hotspot model, we applied multiple diagnostic and robustness assessment approaches. **MCMC convergence diagnostics indicated** that posterior sampling was generally stable: the maximum 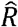 was 1.017, no parameter showed 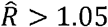 the minimum bulk ESS was 1046, the minimum tail ESS was 1847, and no divergent transitions were observed. These findings suggest overall satisfactory convergence, although a small number of parameters were slightly above 1.01, with no clear evidence of problematic behavior. **Posterior predictive checks (PPCs) indicated** good overall agreement between the predicted and observed data: approximately 92.3% of the observed values fell within the 90% posterior predictive intervals, and the correlation between the observed values and posterior predictive means was high, suggesting that the model reproduced the overall distribution and level of the data reasonably well. **LOO and WAIC results indicated** acceptable model fit and predictive adequacy; however, minor warnings were noted in the LOO/WAIC calculations, with only one observation having a Pareto *k* value in the 0.70–1.00 range, while all remaining observations were within the good range. This suggests that the overall results were broadly robust, although a very small number of potentially influential observations should be noted. **Prior sensitivity analysis indicated** that the main spatial pattern remained highly consistent across different prior specifications: the Spearman correlations between the alternative-prior results and the baseline results were all close to 1, and the top 10 hotspot countries were identical, indicating that the model conclusions were insensitive to prior specification and were therefore robust. More detailed diagnostic results are provided in Text 2 supporting information.

### 3.3. Shared hotspot patterns across the three diseases

#### 3.3.1. Global distribution of the shared hotspot score

The shared hotspot score showed substantial cross-country heterogeneity across the 204-country study panel, indicating that the degree of common hotspot co-occurrence varied widely worldwide. Overall, the score was centred close to zero (mean = 0.000; median = 0.045), but showed a broad distribution, ranging from −5.840177 to 1.275347, with an interquartile range from −0.419 to 0.603. In total, 107 countries had positive shared hotspot scores, whereas 97 had negative values, indicating broad variation in the magnitude and direction of shared hotspot co-occurrence across countries. The highest scores were observed in Eritrea (1.275347), Marshall Islands (1.134881), Burundi (1.122891), Taiwan (1.058625), and Bangladesh (1.033259), indicating stronger co-occurrence of the three respiratory disease hotspots in these settings. In contrast, the lowest scores were found in Tokelau (−5.840177), Niue (−5.688423), Cook Islands (−4.465824), Canada (−1.213604), and Iceland (−0.994190).

The global spatial distribution of the shared hotspot score is presented in Fig 3. Details are provided in S1 Table.

**Fig 3.**
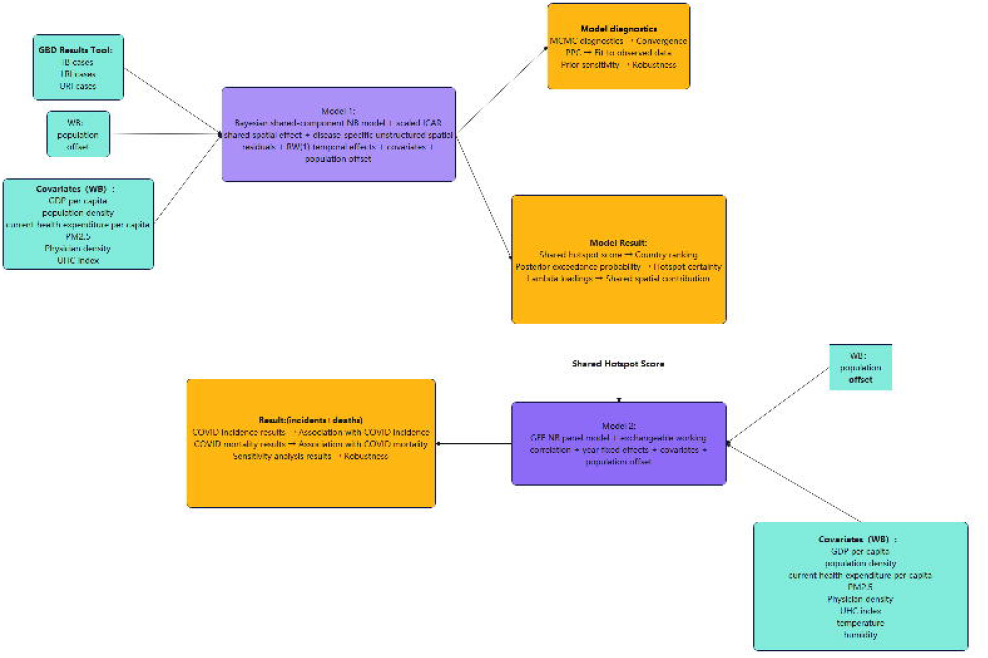
Global distribution of the shared hotspot score across the 204-country study panel. (Note: Map lines delineate study areas and do not necessarily depict accepted national boundaries.)

#### 3.3.2. Posterior exceedance probability of the shared spatial component

Posterior exceedance probabilities of the shared spatial component were estimated as *P*(*S*_*i*_ > 0 | data), providing a probabilistic summary of whether each country exhibited a positive shared spatial effect. A value close to 1 indicates strong posterior support for positive shared hotspot co-occurrence, whereas a value close to 0 indicates weak support; values near 0.5 suggest substantial uncertainty. The resulting map showed marked spatial heterogeneity (refer to Fig 4). High exceedance probabilities were concentrated in much of sub-Saharan Africa and parts of South and Southeast Asia, while low probabilities were observed in North America, Oceania, and several countries in Europe and southern South America. These findings indicate that posterior support for positive shared spatial clustering was strongest in Africa and parts of Asia, but much weaker in several high-income regions. Thus, posterior exceedance probabilities provided an uncertainty-aware complement to the shared hotspot score. Details are provided in Supporting information **Error! Reference source not found**..

**Fig 4.**
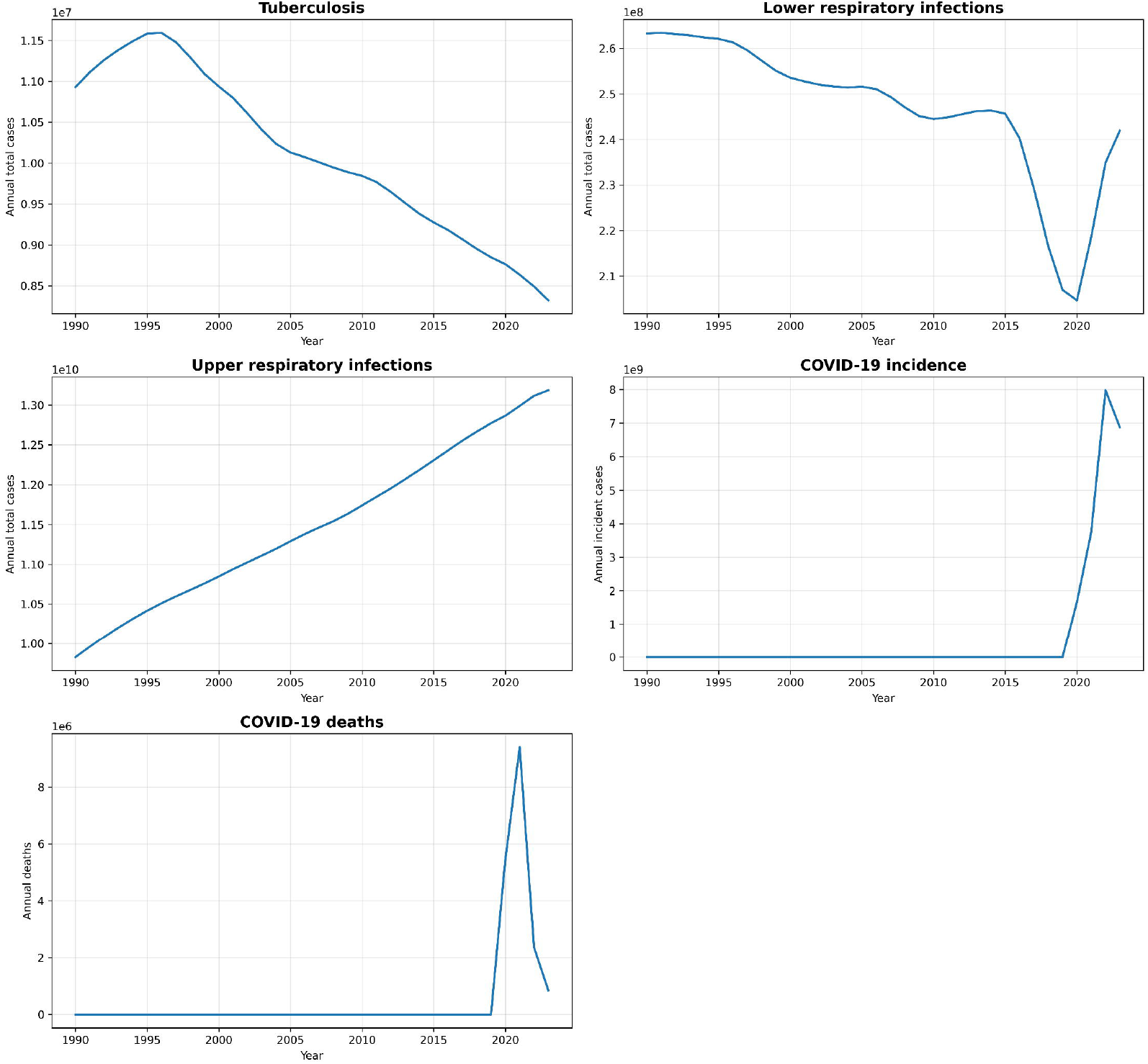
Posterior exceedance probabilities of the shared spatial component. (Note: Map lines delineate study areas and do not necessarily depict accepted national boundaries.)

### 3.4. Contribution of each disease to the shared spatial component

As shown in Fig 5, all three disease-specific shared loadings were positive and their 95% HDIs excluded zero, indicating that lower respiratory infections, tuberculosis, and upper respiratory infections all contributed to the shared spatial component. Tuberculosis had the largest loading (*λ* = 1.657, 95% HDI: 0.883–2.506), followed by lower respiratory infections (*λ* = 1.023, 95% HDI: 0.555–1.569) and upper respiratory infections (*λ* = 0.744, 95% HDI: 0.389–1.144), suggesting that the common spatial pattern was most strongly expressed in tuberculosis. However, Fig 6 shows that the disease-specific spatial effects were not negligible and differed across the three diseases, with residual positive and negative spatial deviations still visible after accounting for the shared component. This indicates that the observed spatial distribution was shaped by both a common shared spatial structure and additional disease-specific spatial heterogeneity.

**Fig 5.**
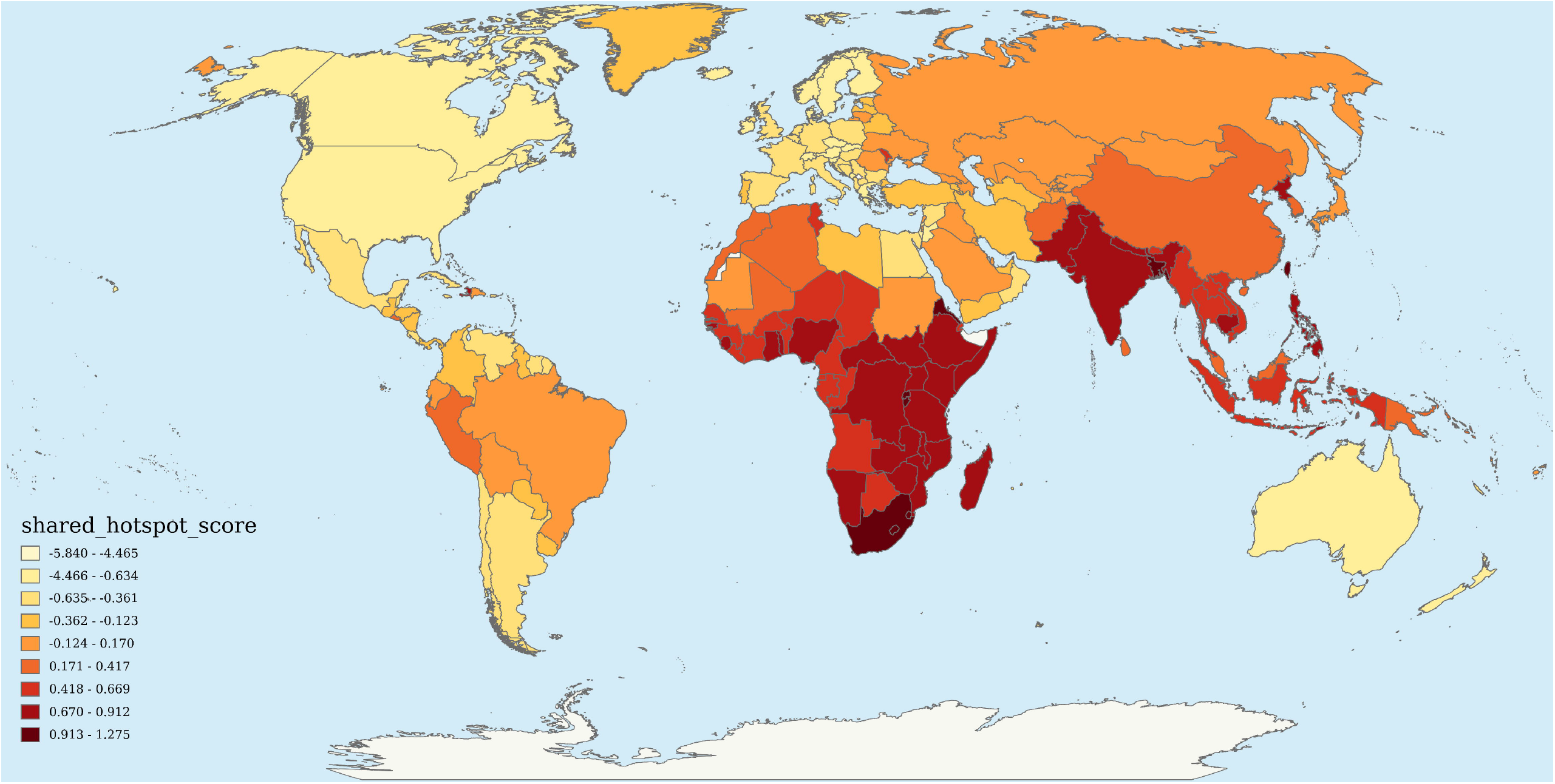
Disease-specific shared loadings for the shared spatial component

**Fig 6.**
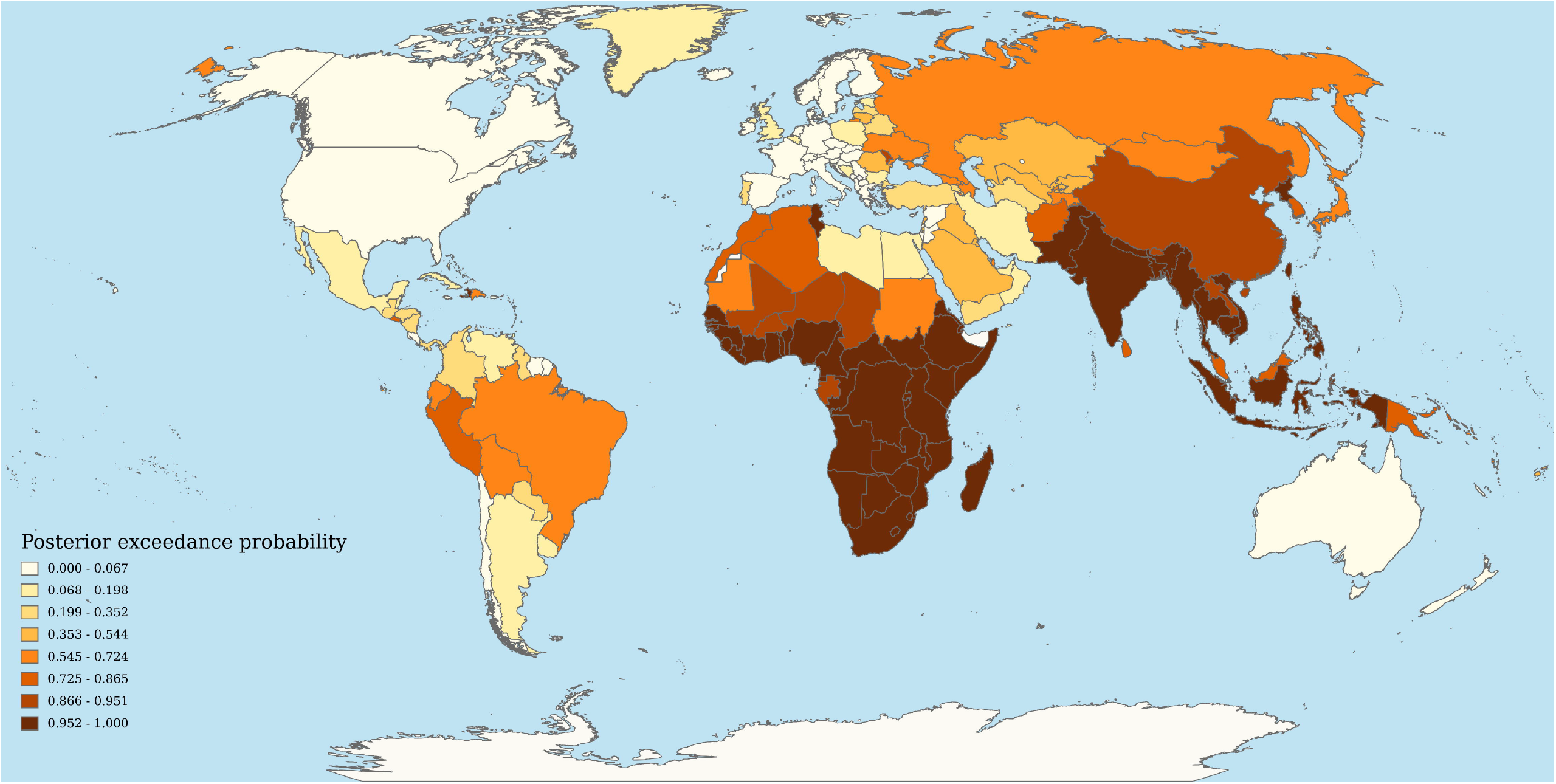
Disease-specific spatial effects after accounting for the shared spatial component. (Note: Map lines delineate study areas and do not necessarily depict accepted national boundaries.)

### 3.5. External validation: associations with COVID-19 incidence and mortality Main continuous-exposure model results

To externally validate the epidemiological relevance of the shared hotspot structure, we examined associations between the standardized shared hotspot score and COVID-19 incidence and mortality using GEE negative binomial models with a population offset and year fixed effects (204 countries, 2020–2023; 816 country-year observations). A one-standard-deviation increase in the shared hotspot score was significantly associated with both higher COVID-19 incidence (IRR = 1.6783, 95% CI: 1.4564–1.9340; P = 8.308 × 10□13) and higher COVID-19 mortality (IRR = 1.7436, 95% CI: 1.5061–2.0186; P = 9.912 × 10□14). These findings provide external support for the epidemiological relevance of the shared hotspot structure identified in the main model.

### 3.6. Sensitivity analysis of the COVID-19 validation models

In the sensitivity analysis, the continuous shared hotspot score was replaced by a binary high-versus-low co-occurrence grouping variable (see Supplementary Text 1 for classification details). Unlike the main continuous-exposure models, the grouped analysis did not show statistically significant associations for either COVID-19 incidence or mortality. The IRR for the high-co-occurrence group relative to the low-co-occurrence group was 0.8341 (95% CI: 0.6163–1.1287; P = 0.2397) for incidence and 0.8760 (95% CI: 0.5579–1.3753; P = 0.5650) for mortality. These null and directionally inconsistent estimates likely reflect information loss and reduced stability after dichotomizing a continuous exposure, rather than robust evidence against the main continuous-score results.

## 4. Discussion

### 4.1. Main findings

This study identified a clear shared spatial hotspot structure across lower respiratory infections, tuberculosis, and upper respiratory infections at the global level. The shared hotspot score showed substantial cross-country heterogeneity, and posterior exceedance probabilities further indicated marked geographic differences in the posterior support for positive shared spatial effects. The countries with the highest shared hotspot scores closely overlapped with those showing the strongest posterior support for positive shared spatial effects. Disease-specific shared loadings showed that all three diseases contributed positively to the shared spatial component, with tuberculosis contributing most strongly. External validation analyses further showed that higher shared hotspot scores were significantly associated with both higher COVID-19 incidence and higher COVID-19 mortality. Together, these findings suggest that the inferred shared hotspot structure reflected a geographically meaningful pattern of common respiratory vulnerability.

### 4.2. Interpretation

The shared spatial component can be interpreted as a latent geographic structure jointly expressed across multiple common respiratory infectious diseases, rather than the spatial signature of any single outcome. This suggests that some countries may face a broad underlying vulnerability that is not disease-specific, but instead reflects common structural conditions affecting respiratory infectious disease transmission and control. Such conditions may include baseline surveillance capacity, access to health care, environmental and social exposures, population mixing, and the resilience of health systems. This interpretation is also broadly consistent with WHO observations in the tuberculosis field, which note that in some high-burden settings, weak routine notification systems, limited vital registration capacity, and lower universal health coverage are associated with difficulty in reliably measuring and controlling TB burden[26]. The stronger shared loading for tuberculosis may indicate that the common spatial pattern is more strongly captured by diseases with sustained background transmission and greater dependence on long-term health system capacity, whereas upper respiratory infections may be more affected by additional disease-specific influences.

### 4.3. Public health implications

The findings have clear public health implications. Countries with high shared hotspot scores appear to bear a heavier background burden of co-occurring common respiratory infectious diseases, and the external validation results suggest that these same settings may also be more vulnerable during large epidemic shocks such as COVID-19. In large-scale respiratory epidemics, countries with stronger economic resources, surveillance systems, and public health response capacity may be better positioned to control transmission earlier, whereas countries facing persistent socioeconomic constraints, weaker health-care infrastructure, and limited surveillance capacity may experience more prolonged or recurrent respiratory disease burden.

Such unresolved vulnerability is not only a national concern, but may also delay global epidemic control by allowing transmission risks to persist across interconnected regions. This indicates that the shared hotspot score may serve as a useful preparedness-oriented indicator, helping to identify settings where routine respiratory disease burden signals broader structural weakness before the next major respiratory epidemic occurs. Rather than treating common respiratory infections and pandemic preparedness as separate agendas, the present results suggest that weaknesses observed under routine conditions may signal broader vulnerability under crisis conditions. From a policy perspective, this supports more integrated and equity-oriented resource allocation strategies that prioritise countries with elevated shared hotspot intensity, particularly through earlier investment in surveillance strengthening, routine respiratory disease control, health-system resilience, and reserve public health response capacity.

### 4.4. Strengths and limitations

This study has several strengths. First, it moved beyond single-disease mapping by identifying a shared spatial component across three common respiratory infectious diseases, thereby capturing common geographic vulnerability rather than disease-specific patterns alone. Second, the Bayesian multivariate spatiotemporal framework allowed the decomposition of disease geography into shared and disease-specific components while accounting for temporal variation and measured covariates. Third, the model was supported by multiple diagnostic assessments, including convergence diagnostics, posterior predictive checks, information-criterion-based summaries, and prior sensitivity analysis. Fourth, the external validation analyses strengthened the interpretability of the shared hotspot score by showing its association with COVID-19 outcomes.

Several limitations should also be noted. First, this was an ecological epidemiological analysis conducted at the country-year level; therefore, the findings should not be interpreted as individual-level or causal associations. Second, the covariates were included as disease-specific adjustment variables and did not directly explain the shared spatial component; therefore, the model cannot identify direct determinants of shared hotspot co-occurrence. Third, the spatial analyses were conducted at the national scale, which may mask important within-country heterogeneity. Fourth, the external validation models supported the epidemiological relevance of the shared hotspot score when analysed as a continuous exposure, whereas the grouped sensitivity analysis provided weaker evidence, suggesting that some findings may depend on exposure specification.

### 4.5. Conclusion

In conclusion, this study identified a geographically structured pattern of shared respiratory vulnerability across tuberculosis, lower respiratory infections, and upper respiratory infections at the global level, concentrated particularly in sub-Saharan Africa, South Asia, and Southeast Asia. External validation confirmed that this shared hotspot structure was significantly associated with both higher COVID-19 incidence and mortality, supporting its epidemiological relevance beyond the diseases used to construct it. These findings suggest that the shared hotspot score may serve as a useful empirical indicator for identifying countries that warrant prioritised attention in preparedness planning and resource allocation ahead of future respiratory epidemics.

## Data Availability

The data used in this study were obtained from publicly available, aggregated, and de-identified sources. The original data sources are described in the manuscript. The processed data and related analysis materials are currently stored in a Google Drive folder and are available at the link provided below.

https://docs.google.com/document/d/1QhDhjhZrwmXf7w08QQr0e9U-_mEWapCT/edit

## Declaration of generative AI use

During the preparation of this work the authors used ChatGPT for translation and language refinement, and Claude in order to assist with statistical model code generation. After using these tools, the authors reviewed and edited the content as needed and take full responsibility for the content of the published article.

## Data availability statement

At the time of submission, the processed data and code can be made available through a third-party <u>anonymous storage link</u>. Upon acceptance, they will be deposited in a public repository and the final link will be provided.

## Acknowledgements

The authors would like to thank Dr. Aisah for her guidance and support throughout this study. The authors also acknowledge the Institute for Health Metrics and Evaluation for providing access to the Global Burden of Disease (GBD) data, and the World Bank for making the covariate data publicly available.

