## Supplementary Text S1 for "Shared respiratory infectious disease hotspots identify priority countries for pandemic preparedness: a Bayesian spatiotemporal analysis with COVID-19 external validation"

**Supplementary Text 1: Detailed Model Specification**

### 1. Main Model (Model 1): Bayesian Multivariate Shared-Component Spatiotemporal Model

#### 1.1. Overview

To identify cross-disease shared hotspot patterns while allowing disease-specific spatial and temporal heterogeneity, we implemented a Bayesian multivariate shared-component spatiotemporal model for three respiratory diseases: tuberculosis (TB), lower respiratory infections (LRI), and upper respiratory infections (URI) (1, (2). The model was fitted to annual country-level incidence counts from 204 countries over 1990–2023, yielding 20,808 total observations (204 countries × 3 diseases × 34 years).

The core idea of the shared-component framework is to decompose the spatial variation of multiple related diseases into two parts: (i) a shared spatial component that captures the common geographic pattern jointly expressed across all three diseases, and (ii) disease-specific spatial deviations that capture the residual spatial variation unique to each disease. By separating these two sources of spatial variation, the model can identify countries that are simultaneously elevated across all three diseases (shared hotspots), as distinct from countries where only one disease is elevated (disease-specific patterns).

#### 1.2. Observation model

Let $y_{ikt}$ denote the observed incidence count for country $i$ ($i=1,\ldots,204$), disease $k$ ($k=1,2,3$, corresponding to TB, LRI, URI), and year $t$ ($t=1990,\ldots,2023$). The observation model was specified as:

$$y_{ikt}\sim\text{NegBin}\left( \mu_{ikt},\phi_{k} \right)$$

where $\mu_{ikt}$ is the expected count and $\phi_{k}$ is the disease-specific overdispersion (or concentration) parameter of the negative binomial distribution. A negative binomial distribution was used instead of a Poisson distribution because it accommodates overdispersion — that is, the common situation in which the observed variance in disease counts exceeds the mean — by introducing the additional parameter $\phi_{k}$ (3).Larger values of $\phi_{k}$ indicate less overdispersion (approaching a Poisson distribution), while smaller values indicate greater overdispersion. Each disease was assigned its own $\phi_{k}$, allowing the degree of overdispersion to differ across TB, LRI, and URI.

**Linear predictor**

The log-expected count was modeled as a linear combination of six additive components:

$$log\left( \mu_{ikt} \right)=\underset{\text{(a) offset}}{\underset{⏟}{\log\left( P_{it} \right)}}+\underset{\text{(b) intercept}}{\underset{⏟}{\alpha_{k}}}+\underset{\text{(c) covariates}}{\underset{⏟}{\sum_{p=1}^{P} \beta_{kp}X_{itp}}}+\underset{\text{(d) shared spatial}}{\underset{⏟}{\lambda_{k} s_{i}}}+\underset{\text{(e) disease-specific spatial}}{\underset{⏟}{u_{ki}}}+\underset{\text{(f) temporal}}{\underset{⏟}{\theta_{kt}}}$$

Each component is described in detail below.

**(a) Population offset:** $\mathbf{log}\left( \boldsymbol{P}_{\boldsymbol{it}} \right)$

$P_{it}$ denotes the total population of country $i$ in year $t$. Including $\log\left( P_{it} \right)$ as an offset (a fixed term with coefficient 1) on the log scale means that the model effectively estimates incidence rates (counts per population) rather than raw counts. This adjustment is necessary because countries with larger populations are expected to have more cases simply due to having more people at risk, regardless of underlying disease risk. After accounting for the offset, all remaining terms in the linear predictor reflect differences in log-rate rather than log-count.

(**b) Disease-specific intercept:** $\boldsymbol{\alpha}_{\boldsymbol{k}}$

$\alpha_{k}$ represents the baseline log-rate for disease $k$, averaged across all countries and years after accounting for other model components. Since the three diseases differ greatly in their overall incidence levels (e.g., URI is far more common than TB), each disease requires its own intercept. There are three intercept parameters in total ($\alpha_{\text{TB}}$, $\alpha_{\text{LRI}}$, $\alpha_{\text{URI}}$).

**(c) Covariate effects:** $\sum_{\boldsymbol{p}\mathbf{=}\boldsymbol{1}}^{\boldsymbol{P}} \boldsymbol{\beta}_{\boldsymbol{kp}}\boldsymbol{X}_{\boldsymbol{itp}}$

$X_{itp}$ denotes the value of the $p$-th covariate for country $i$ in year $t$, and $\beta_{kp}$ is the corresponding disease-specific regression coefficient. The six covariates included were: gross domestic product (GDP) per capita, population density, current health expenditure per capita, ambient PM₂.₅ concentration, the Universal Health Coverage (UHC) service coverage index, and physician density.

Prior to model fitting, GDP per capita, population density, health expenditure per capita, and PM₂.₅ were natural log-transformed ($\log_{1+x}$) to reduce right skewness. All six covariates were then standardized to zero mean and unit variance across the full panel. The regression coefficients $\beta_{kp}$ are disease-specific, meaning that each covariate is allowed to have a different effect on each of the three diseases. For example, GDP per capita may have a stronger protective association with TB than with URI. There are $3\times6=18$ covariate coefficients in total.

The role of the covariates in this model is to adjust for known country-level risk factors, so that the shared spatial component $s_{i}$ captures geographic co-elevation that cannot be explained by these measured factors alone.

**(d) Shared spatial component:** $\boldsymbol{\lambda}_{\boldsymbol{k}} \boldsymbol{s}_{\boldsymbol{i}}$

This is the core component of the shared-component framework and consists of two elements:

**The shared spatial effect** $s_{i}$ is a single latent value assigned to each country, shared identically across all three diseases. It represents the common geographic risk pattern that is jointly expressed by TB, LRI, and URI. A country with a high $s_{i}$ exhibits higher-than-expected incidence across all three diseases (after adjusting for covariates and temporal trends), while a country with a low $s_{i}$ exhibits lower-than-expected incidence. Crucially, $s_{i}$ is time-invariant — each country has one value of $s_{i}$ across the entire 34-year study period. Temporal variation is handled separately by $\theta_{kt}$.

The spatial structure of $s_{i}$ was modeled using a scaled intrinsic conditional autoregressive (ICAR) prior (4), which encodes the expectation that geographically adjacent countries should have similar values. Formally, the ICAR prior specifies that each country’s $s_{i}$ is conditionally distributed as a normal distribution centered on the mean of its neighbors’ values, with the degree of smoothing controlled by a global standard deviation parameter $\sigma_{s}$. The scaling factor ensures that the marginal variance of $s_{i}$ is comparable across spatial graphs with different topologies [6]. Countries without shared borders (island nations) were connected to their five nearest neighbors by centroid distance (KNN fallback with $K=5$) to ensure a connected spatial graph. A sum-to-zero constraint was imposed on $s_{i}$ for identifiability, so that the values are interpretable as deviations from the global mean.

For each country, the posterior estimate of $s_{i}$ is jointly determined by two sources of information: (i) the data — specifically, the residual geographic co-elevation of the three diseases after accounting for measured covariates, disease-specific temporal trends, and disease-specific spatial deviations; and (ii) the ICAR prior — which provides spatial smoothing toward neighboring countries. A country receives a high $s_{i}$ when all three diseases show higher-than-expected incidence in that country and its neighbors after adjustment.

**The disease-specific loading** $\lambda_{k}$ scales how strongly each disease $k$ contributes to (or is influenced by) the shared spatial pattern $s_{i}$. A larger $\lambda_{k}$ means that disease $k$ is more strongly associated with the common geographic pattern. All $\lambda_{k}$ values were constrained to be non-negative via a half-normal prior, which fixes the direction of $s_{i}$ (i.e., positive $s_{i}$ always indicates higher shared risk) and prevents sign-switching across diseases. There are three loading parameters ($\lambda_{\text{TB}}$, $\lambda_{\text{LRI}}$, $\lambda_{\text{URI}}$).

Together, the product $\lambda_{k} s_{i}$ means that country $i$’s contribution from the shared spatial pattern to disease $k$ is proportional to $s_{i}$, but scaled by how strongly disease $k$ participates in the common pattern.

**(e) Disease-specific spatial deviation:** $\boldsymbol{u}_{\boldsymbol{ki}}$

$u_{ki}$ captures the residual spatial variation in disease $k$ for country $i$ that is not explained by the shared spatial component. For example, a country might have an unusually high TB burden due to HIV co-infection — a factor that does not equally affect URI — and this would be absorbed by $u_{\text{TB},i}$ rather than $s_{i}$. The $u_{ki}$ terms were modeled as independent normal random effects:

$$\boldsymbol{u}_{\boldsymbol{ki}}\boldsymbol{\sim}\text{Normal}\left( \boldsymbol{0}\mathbf{,}\boldsymbol{\sigma}_{\text{sd}} \right)$$

where $\sigma_{\text{sd}}$ is a shared standard deviation across all diseases. There are $3\times204=612$ disease-specific spatial deviation parameters.

By including both $\lambda_{k}s_{i}$ and $u_{ki}$, the model decomposes each country’s total spatial effect for disease $k$ into a shared part ($\lambda_{k}s_{i}$, common across diseases) and a disease-specific part ($u_{ki}$, unique to each disease).

**(f) Disease-specific temporal effect:** $\boldsymbol{\theta}_{\boldsymbol{kt}}$

$\theta_{kt}$ captures year-to-year variation in disease $k$ that is common across all countries. For example, the global decline in TB incidence over recent decades, or seasonal fluctuations in URI, would be captured by $\theta_{kt}$. These temporal effects were modeled using a first-order random walk [RW(1)] prior (5):

$$\boldsymbol{\theta}_{\boldsymbol{k}\mathbf{,}\boldsymbol{t}\mathbf{+}\boldsymbol{1}}\mathbf{=}\boldsymbol{\theta}_{\boldsymbol{k}\mathbf{,}\boldsymbol{t}}\mathbf{+}\boldsymbol{\epsilon}_{\boldsymbol{kt}}\mathbf{,} \boldsymbol{\epsilon}_{\boldsymbol{kt}}\boldsymbol{\sim}\text{Normal}\left( \boldsymbol{0}\mathbf{,}\boldsymbol{\sigma}_{\text{rw}\mathbf{,}\boldsymbol{k}} \right)$$

where $\sigma_{\text{rw},k}$ controls the smoothness of the temporal trajectory for disease $k$. A smaller $\sigma_{\text{rw},k}$ produces smoother (more slowly changing) temporal effects, while a larger value allows more rapid year-to-year fluctuations. Each disease has its own $\sigma_{\text{rw},k}$, allowing different degrees of temporal variability across diseases. The temporal effects were centered (i.e., $\sum_{t} \theta_{kt}=0$ for each $k$) for identifiability, so that the overall level is absorbed by the intercept $\alpha_{k}$. There are $3\times34=102$ temporal effect parameters.

Note that $\theta_{kt}$ varies by disease and year but not by country. This means the model assumes that temporal trends are shared across all countries within each disease. Country-specific temporal deviations are absorbed into the negative binomial residual variance.

#### 1.3. Summary of model dimensions

| Component | Notation | Dimensions | Total parameters |
| --- | --- | --- | --- |
| Population offset | $\log\left( P_{it} \right)$ | 204 × 34 | Fixed (data) |
| Disease intercept | $\alpha_{k}$ | 3 | 3 |
| Covariate effects | $\beta_{kp}$ | 3 × 6 | 18 |
| Shared spatial | $s_{i}$ | 204 | 204 |
| Shared loading | $\lambda_{k}$ | 3 | 3 |
| Disease-specific spatial | $u_{ki}$ | 3 × 204 | 612 |
| Temporal effects | $\theta_{kt}$ | 3 × 34 | 102 |
| Overdispersion | $\phi_{k}$ | 3 | 3 |
| Variance parameters | $\sigma_{s},\sigma_{\text{sd}},\sigma_{\text{rw},k}$ | 1 + 1 + 3 | 5 |

#### 1.4. Prior distributions

**Prior distributions**

Most prior distributions in the model were specified as weakly informative priors because strong quantitative prior information was unavailable (6). Following the general principle of Gelman (2006), we used proper weakly informative priors to constrain parameters within epidemiologically plausible ranges while allowing the likelihood to dominate posterior inference (7). These weakly informative priors were applied to the disease-specific intercepts, standardized covariate coefficients, shared spatial scale, shared-component loadings, and disease-specific spatial scale.

The numerical prior scales were selected according to the scale and interpretation of each model component rather than being treated as fixed values directly prescribed by previous literature. To evaluate whether these choices affected substantive inference, we conducted a systematic one-at-a-time prior sensitivity analysis using more diffuse alternatives. The sensitivity results showed that scale-only widening of the weakly informative priors produced highly stable posterior conclusions, with nearly identical country-level shared-hotspot rankings and unchanged Top-10 hotspot country lists. A limited degree of sensitivity was observed only when the disease-specific iid spatial component was assigned a heavier-tailed Student-t prior rather than a Gaussian prior. Even under this alternative specification, the overall ranking structure and broad spatial pattern were largely preserved. Therefore, the main hotspot rankings were robust to the numerical scales of the weakly informative priors, with only limited sensitivity to the distributional form of the disease-specific spatial component.

Complete prior specifications and justifications are summarized in Table 1.

Table 1 Prior distributions and justification

| **Parameter** | **Prior** | **Prior type** | **Justification** |
| --- | --- | --- | --- |
| α_k, disease-specific intercepts | Normal(0, 2) | Weakly informative | Used because strong prior information on baseline log-rates was unavailable; allows a broad range of disease-specific baseline risks. |
| β_kp, standardized covariate coefficients | Normal(0, 1) | Weakly informative (8) | Applied to standardized covariates; allows substantial per-standard-deviation log-rate effects while limiting implausibly extreme coefficient estimates. |
| s_i, shared spatial component | Scaled ICAR | Structural spatial prior | Uses the scaled ICAR formulation to ensure interpretable spatial scaling across adjacency graphs. |
| σ_s, shared spatial scale | HalfNormal(1) | Weakly informative | Prior placed on the standard-deviation scale; provides weak regularization for the magnitude of the shared spatial component. |
| λ_k, shared-component loadings | HalfNormal(1) | Weakly informative | Non-negative loading prior prevents sign switching of the shared spatial component and weakly regularizes disease-specific contributions to the shared component. |
| u_ki, disease-specific iid spatial effects | Normal(0, σ_sd) | Weakly informative hierarchical prior | Centres disease-specific spatial deviations at zero, with their magnitude governed by σ_sd. |
| σ_sd, disease-specific spatial scale | HalfNormal(1) | Weakly informative | Prior placed on the standard-deviation scale; allows disease-specific spatial variation while providing hierarchical shrinkage. |
| θ_kt, temporal effect | RW(1), σ_rw,k ~ HalfNormal(0.5) | Mildly regularizing | Encourages temporal smoothness on the annual log-rate scale while allowing year-to-year changes when supported by the data. |
| φ_k, negative binomial overdispersion | Gamma(2, 0.5) | Mildly regularizing | Places prior mass around moderate overdispersion while down-weighting, but not excluding, very small or very large dispersion values. |

A comprehensive prior sensitivity analysis was conducted under more diffuse alternative specifications, including Normal(0, 5) for α_k, Normal(0, 2) for β_kp, and HalfNormal(2) for σ_s, λ_k, and σ_sd. These scale-only modifications produced highly consistent country-level shared-hotspot rankings and unchanged Top-10 hotspot country lists relative to the baseline model, supporting the interpretation that the main conclusions were not materially driven by the weakly informative prior scales. Detailed sensitivity results are reported in Supplementary Text S2.

#### 1.5. Bayesian posterior inference

The observation model (negative binomial likelihood) and the prior distributions specified above jointly define the full Bayesian model. Inference proceeds by combining the likelihood with the priors via Bayes’ theorem to obtain the joint posterior distribution:

$$\begin{aligned} p\left( \alpha, \beta, \lambda, s, u, \theta, \phi, \sigma\mid y \right) \\ \propto\prod_{i,k,t} p\left( y_{ikt} \mid\mu_{ikt}, \phi_{k} \right) \times\pi\left( \alpha\right) \times\pi\left( \beta\right) \times\pi\left( \lambda\right) \times\pi\left( s \mid\sigma_{s} \right) \\ \times\pi\left( u \mid\sigma_{sd} \right) \times\pi\left( \theta\mid\sigma_{rw} \right) \times\pi\left( \phi\right) \times\pi\left( \sigma_{s}, \sigma_{sd}, \sigma_{rw} \right) \end{aligned}$$

where the left-hand side is the joint posterior over all model parameters and latent effects, the product term is the negative binomial likelihood across all 20,808 country–disease–year observations, and π(·) denotes the prior distributions specified above. Because this joint posterior does not have a closed-form solution, posterior samples were obtained via Markov chain Monte Carlo (MCMC) sampling, as described in Section 1.7.

#### 1.6. Derivation of the shared hotspot score

The shared hotspot score $H_{i}$for each country was derived from the posterior output as a loading-weighted summary:

$$H_{i}=\frac{\sum_{k} \bar{\lambda}_{k}\cdot\bar{s}_{i}}{\sum_{k} \bar{\lambda}_{k}}$$

where $\bar{\lambda}_{k}$and $\bar{s}_{i}$denote the posterior means of the disease-specific loading and the shared spatial component, respectively. Because $s_{i}$does not vary across diseases, this expression reduces to $H_{i}=\bar{s}_{i}$. In practice, $\bar{s}_{i}$was computed as the mean of 32,000 posterior samples (16 chains × 2,000 post-warmup draws). Before entering the external validation model (Model 2), $H_{i}$ was standardized to zero mean and unit variance across the 204 countries, so that the resulting incidence rate ratios are interpretable as the effect per one-standard-deviation increase.

Positive values of $H_{i}$ indicate countries whose respiratory disease geography is consistent with a shared hotspot pattern — that is, all three diseases are jointly elevated after adjustment. Negative values indicate departure from the shared pattern. The posterior exceedance probability $P\left( s_{i}>0\mid\text{data} \right)$ provides an uncertainty-aware complement: values close to 1 indicate strong posterior support for positive shared spatial co-occurrence, values close to 0 indicate the opposite, and values near 0.5 indicate substantial uncertainty.

#### 1.7. Posterior inference and computational details

Posterior inference was conducted using MCMC sampling implemented in PyMC with the No-U-Turn Sampler (NUTS). The sampling configuration was as follows: 16 independent chains, each with 2,000 warmup (tuning) iterations and 2,000 post-warmup draws, yielding 32,000 posterior samples in total. The target acceptance rate was set to 0.95.

Convergence was assessed using: - Trace plots for visual inspection of chain mixing - The Gelman–Rubin convergence statistic ($\hat{R}$), with $\hat{R}<1.05$ as the convergence threshold - Effective sample size (ESS), with ESS > 400 as the minimum threshold for reliable posterior summaries - The number of divergent transitions, with zero divergences as the target

Model adequacy was assessed using: - Posterior predictive checks (PPCs), comparing the distribution of observed data with data simulated from the posterior predictive distribution - Leave-one-out cross-validation (LOO) and the widely applicable information criterion (WAIC) as summaries of out-of-sample predictive performance - Pareto $k$ diagnostics for identifying influential observations

Additional robustness assessments included prior sensitivity analysis (described above) and correlation of the shared hotspot score with the Global Health Security Index (GHSI) 2021 overall score.

### 2. External Validation Model (Model 2): GEE Negative Binomial

#### 2.1. Model description

Annual country-level COVID-19 incidence counts and COVID-19 death counts were analyzed separately using generalized estimating equations (GEE) with a negative binomial distribution, a log link, an exchangeable working correlation structure, and a log-population offset (9). A negative binomial distribution was used instead of a Poisson distribution because overdispersion was expected in annual country-level COVID-19 count data (3).

The model was specified as:

$$Y_{it}\sim\text{NegBin}\left( \mu_{it},\phi\right)$$

$$\log\left( \mu_{it} \right)=log\left( P_{it} \right)+\beta_{0}+\beta_{1}H_{i}+\sum_{m=1}^{M} \gamma_{m}Z_{mit}+\delta_{t}$$

where:

$Y_{it}$ is the annual COVID-19 count (incidence or deaths) for country $i$ in year $t$

$P_{it}$ is the population offset

$H_{i}$ is the standardized shared hotspot score (derived from Model 1)

$\gamma_{m}$is its corresponding coefficient

$Z_{mit}$ denotes the $m$-th time-varying covariate

$\delta_{t}$ represents year fixed effects (2020–2023)

$\beta_{1}$ is the coefficient of primary interest: it quantifies the association between the shared hotspot score and COVID-19 outcomes, expressed as an incidence rate ratio (IRR = $\exp\left( \beta_{1} \right)$) per one-standard-deviation increase in $H_{i}$

Year fixed effects $\delta_{t}$ were included to account for the global pandemic trajectory (e.g., the emergence of variants, vaccine rollout). Country fixed effects were not included because $H_{i}$ is time-invariant at the country level and would be collinear with country fixed effects.

The covariates comprised GDP per capita, population density, health expenditure per capita, PM₂.₅, physician density, UHC index, July temperature, and July humidity. All continuous covariates were standardized before model fitting, with GDP per capita, population density, health expenditure per capita, PM₂.₅, and physician density log-transformed beforehand to reduce skewness.

#### 2.2. Sensitivity analysis

As a sensitivity analysis, an alternative GEE negative binomial model was fitted after classifying countries into high- and low-shared-hotspot groups using quartile-based cutoffs: the top 25% were defined as high, the bottom 25% as low, and the middle 50% were excluded. The continuous shared hotspot score was replaced by a binary group indicator in this model.
