## Supplementary Text S2 for "Shared respiratory infectious disease hotspots identify priority countries for pandemic preparedness: a Bayesian spatiotemporal analysis with COVID-19 external validation"

**Text 2 Model 1 Diagnostic Report**

### 1. MCMC convergence diagnostics

MCMC convergence diagnostics were used to evaluate whether posterior sampling had reached a stable target distribution and whether the Markov chains mixed adequately across iterations. Good convergence is essential to ensure that posterior summaries are reliable and not driven by incomplete exploration of the parameter space.

Convergence was assessed using trace plots, Gelman-Rubin convergence statistics ($\hat{R}$), effective sample sizes (ESS), and the presence or absence of divergent transitions. In general, trace plots with stable horizontal mixing and no systematic drift indicate satisfactory convergence. $\hat{R}$values close to 1 indicate good agreement across chains, whereas values substantially above 1 suggest possible non-convergence. Larger bulk and tail ESS values indicate more reliable estimation of posterior means and tails. The absence of divergent transitions further supports stable sampling.

The MCMC convergence diagnostics indicated generally satisfactory posterior sampling performance. The trace plots of key parameters showed stable trajectories without obvious drift or abrupt shifts, and the posterior density plots were broadly consistent across chains, suggesting acceptable chain mixing. Quantitatively, the maximum $\hat{R}$was 1.017, no parameter showed $\hat{R}>1.05$, the minimum bulk ESS was 1046, and the minimum tail ESS was 1847. In addition, no divergent transitions were observed. Taken together, these findings support overall satisfactory convergence, although a small number of parameters were slightly above 1.01, indicating that convergence was acceptable rather than perfect. Detailed results are provided in Supplementary Figure 1 and Supplementary Table 1.

Table 1 Summary of MCMC convergence diagnostics for Model 1

| maximum $\hat{R}$ | number of parameters with $\hat{R}>1.01$ | number of parameters with $\hat{R}>1.05$ | minimum bulk ESS | number of parameters with bulk ESS < 400 | minimum tail ESS | divergent transitions |
| --- | --- | --- | --- | --- | --- | --- |
| 1.0171 | 69 | 0 | 1046.3302 | 0 | 1846.7949 | 0 |

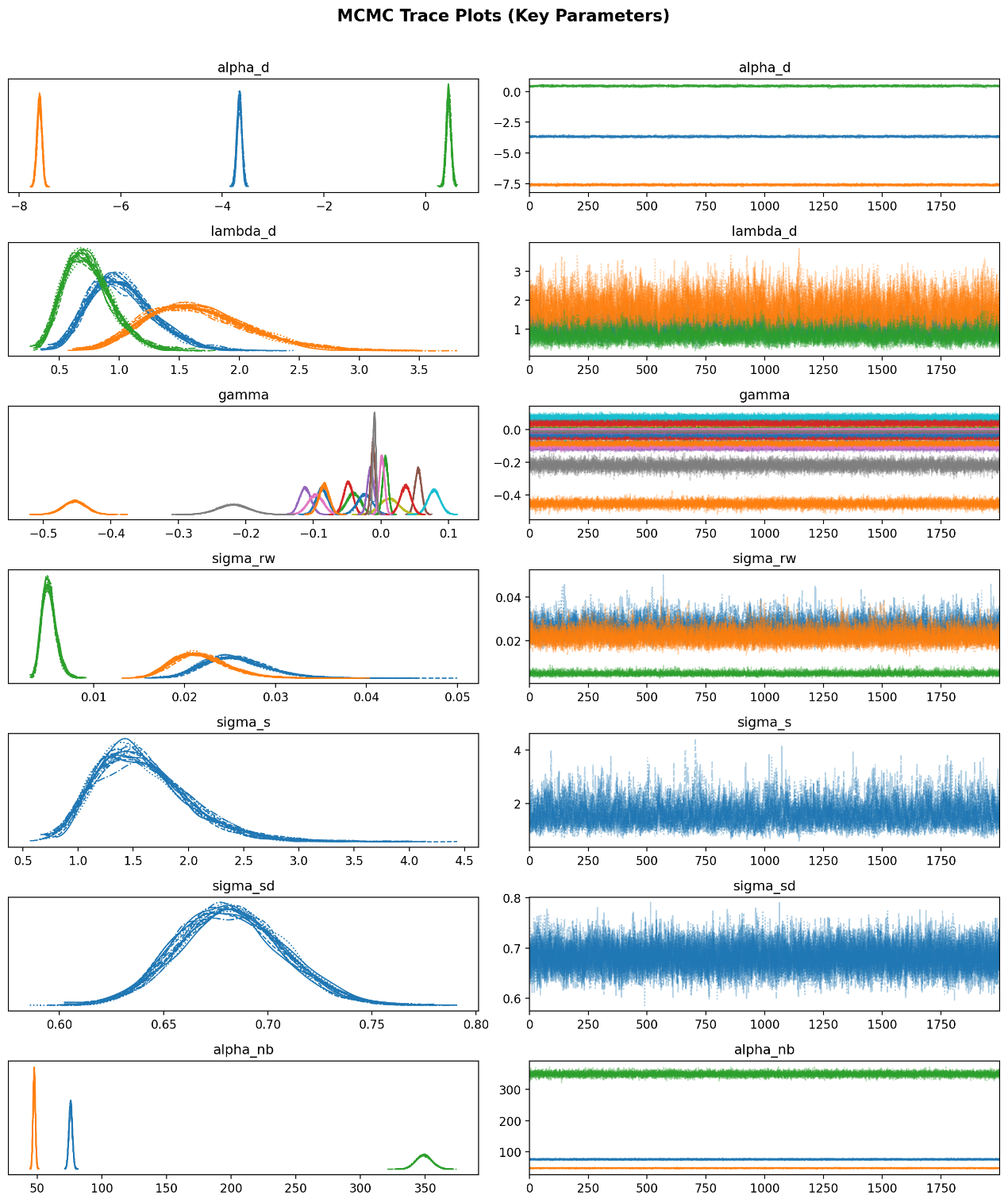

Figure 1 MCMC trace plots and posterior density plots for key parameters in Model 1

Note: In Figure 1,for parameters with shape=3 (alpha_d, alpha_nb, sigma_rw, lambda_d, gamma), each colour represents one disease (TB, LRI, URI) rather than a separate chain.

### 2. Posterior predictive checks (PPCs)

Posterior predictive checks were used to assess whether the fitted model could reproduce the main features of the observed data. This diagnostic evaluates model adequacy by comparing observed values with data simulated from the posterior predictive distribution.

Model fit was considered better when the posterior predictive distributions broadly overlapped the observed distributions, when observed values were well aligned with posterior predictive means, and when most observed values fell within the posterior predictive uncertainty intervals. Marked discrepancies between observed and predicted distributions, systematic deviations from the 45-degree line in observed-versus-predicted plots, or consistently low interval coverage would indicate inadequate model fit.

The posterior predictive checks indicated good overall agreement between the fitted model and the observed data. Approximately 92.3% of the observed values fell within the 90% posterior predictive intervals, suggesting appropriate overall uncertainty calibration. In addition, the correlation between the observed values and the posterior predictive means was high, and the observed-versus-predicted plots showed that most points lay close to the 45-degree reference line. Distribution-based PPCs further showed substantial overlap between the observed and posterior predictive distributions both overall and within each disease. These results indicate that the model reproduced the overall level and distributional structure of the data reasonably well. Detailed results are provided in Figure 2 and Figure 3 and Figure 4.
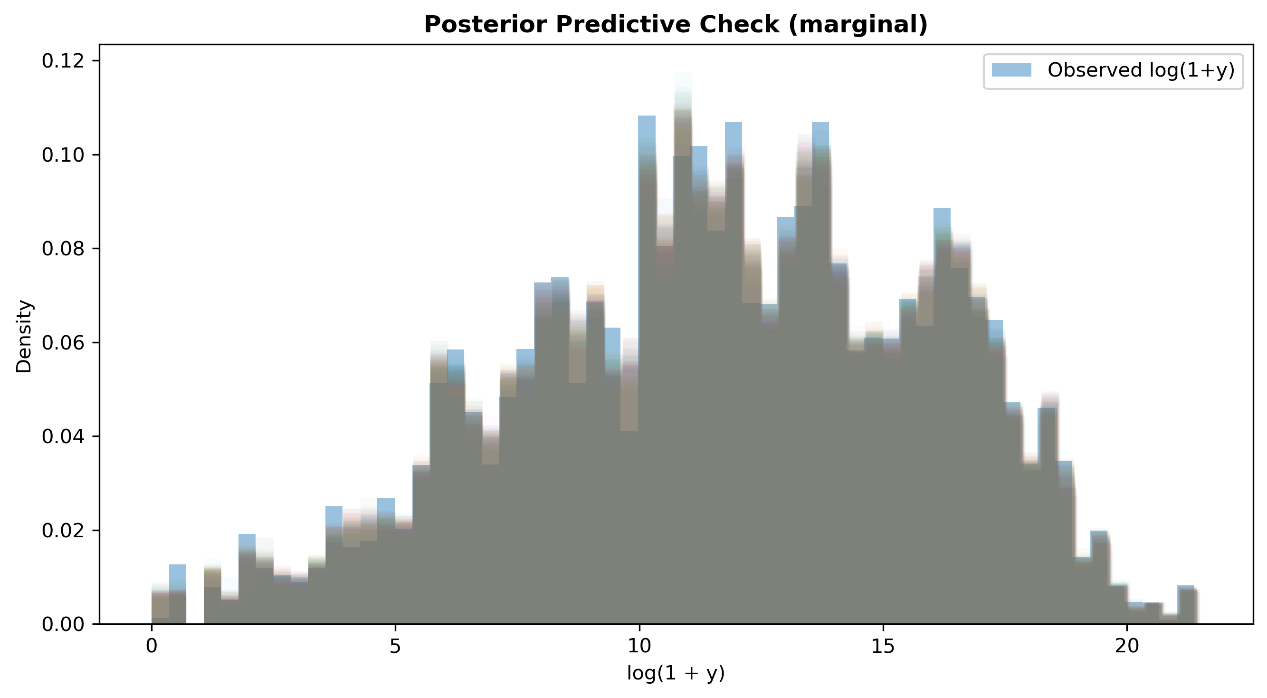

Figure 2 Marginal posterior predictive check

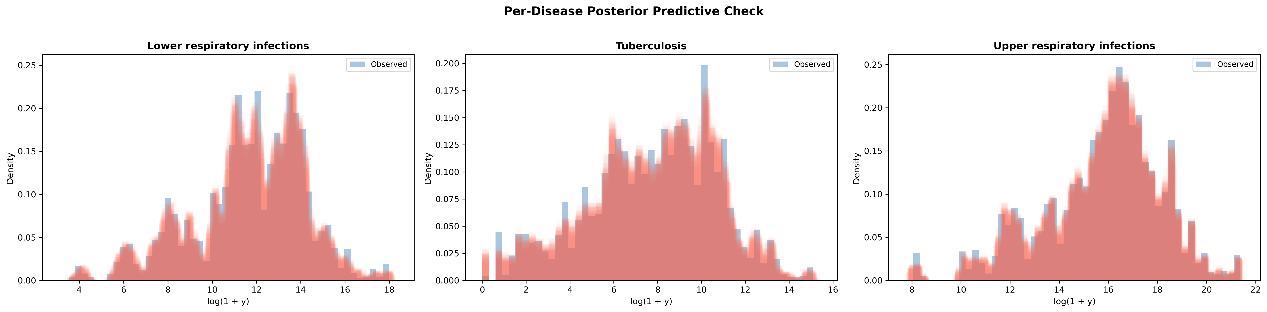

Figure 3 Per-disease posterior predictive checks

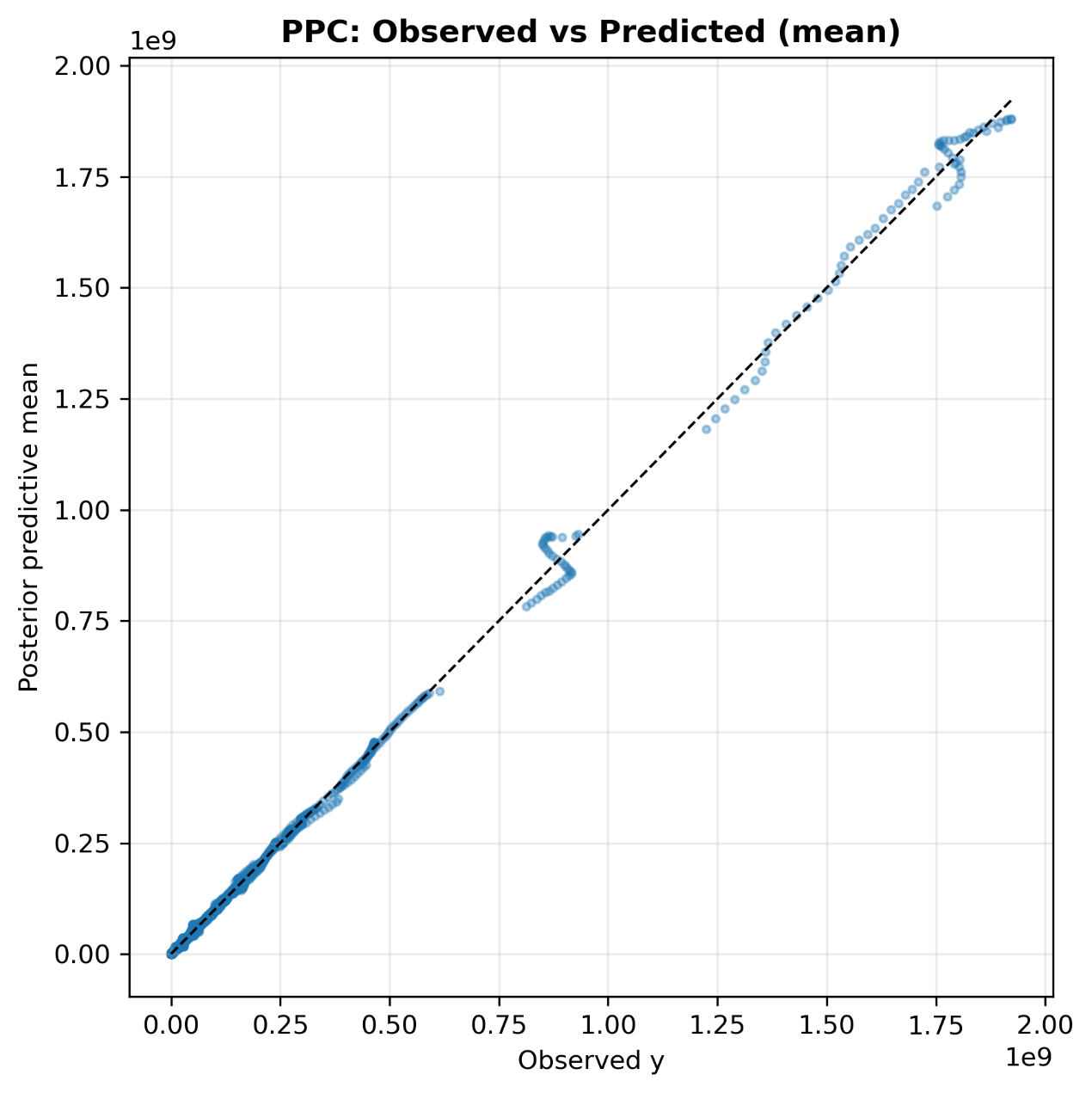

Figure 4 Observed versus posterior predictive mean

### 3. Model Fit: LOO-CV and WAIC

#### 3.1. Method

This study assessed whether the fitted model adequately describes the observed data using two complementary information criteria.

**Leave-one-out cross-validation (LOO-CV)** evaluates predictive accuracy by estimating how well the model would predict each observation if that observation were left out of the fitting process. Rather than re-fitting the model 20,808 times, LOO-CV is efficiently approximated via Pareto-smoothed importance sampling [1]. The key output is the expected log-pointwise-predictive-density ($ELPD_{LOO}$), where higher (less negative) values indicate better out-of-sample predictive performance.

**The widely applicable information criterion (WAIC)** provides a similar estimate of out-of-sample predictive accuracy using a different computational approach [2]. It serves as a cross-check on LOO-CV; substantial disagreement between the two would suggest numerical instability.

**Both criteria also report two important diagnostic quantities:**

**Effective number of parameters (**$p_{eff}$**)**: measures model complexity—the number of parameters the model effectively uses to fit the data. A value much larger than the actual number of explicit parameters indicates that the random effects are capturing substantial variation; a value approaching the number of observations would suggest overfitting.

**Pareto** $k$**diagnostic (LOO-CV only)** [3]: assesses the reliability of the importance sampling approximation for each observation. Each of the 20,808 observations receives a $k$value:

$k\leq0.70$(“good”): the LOO estimate for this observation is reliable;

$0.70<k\leq1.0$(“bad”): the estimate may be unreliable—this observation is unusually influential;

$k>1.0$(“very bad”): the estimate is unreliable and the model may be misspecified for this observation.

A high proportion of “bad” or “very bad” $k$values would indicate that certain observations exert excessive influence on the model, potentially signalling outliers or model misspecification.

#### 3.2. Results

The LOO and WAIC estimates were highly consistent, with near-identical ELPD and standard errors (Table 2), confirming numerical stability. The effective number of parameters ($p_{LOO}=676.50$; $p_{WAIC}=675.23$) is plausible for a model with 204 countries × 3 diseases over 34 years, shared and disease-specific spatial random effects, and disease-specific temporal random walks—indicating that the model uses a moderate degree of flexibility without overfitting ($p_{eff}\ll20,808$ total observations). Critically, 20,807 of 20,808 observations had Pareto $k\leq0.70$(“good”), and only one observation fell within the $0.70<k\leq1.0$range, indicating that the LOO-CV approximation was broadly reliable overall, although a very small number of potentially influential observations should be noted. Consistent with this, both LOO and WAIC produced minor calculation warnings, so these results should be interpreted as supporting acceptable and broadly stable predictive adequacy rather than perfect numerical regularity.

Table 2 PriModel comparison via LOO-CV and WAIC

| Metric | ELPD | SE | p_eff |
| --- | --- | --- | --- |
| LOO-CV | -226075.73 | 582.05 | 676.50 |
| WAIC | -226074.46 | 582.03 | 675.23 |

**Pareto** $k$**diagnostics:**

| Category | Count | Percentage |
| --- | --- | --- |
| $(-\infty,0.70]$(good) | 20,807 | 100.0% |
| $(0.70,1.0]$(bad) | 1 | 0.0% |
| $\left( 1.0 , \infty\right)$(very bad) | 0 | 0.0% |

### 4. Prior sensitivity analysis

Prior sensitivity analysis was used to assess whether the main model conclusions were robust to reasonable alternative prior specifications. This is particularly important in Bayesian hierarchical models, where overly influential priors may distort posterior inference.

Robustness was judged by comparing the spatial patterns obtained under the baseline prior with those obtained under alternative priors. Good robustness is indicated when the spatial ranking of countries remains highly similar across prior settings, when correlation coefficients between baseline and alternative results are close to 1, and when the highest-priority hotspot countries remain largely unchanged. Major reordering of countries or large changes in hotspot patterns would suggest sensitivity to prior assumptions.

The prior sensitivity analysis indicated strong robustness of the inferred shared hotspot structure. Across the alternative prior specifications, the main spatial pattern remained highly consistent with the baseline results. The Spearman correlations between the alternative-prior outputs and the baseline outputs were all close to 1, and the top 10 hotspot countries were identical across prior settings. These findings indicate that the substantive conclusions of the model were not materially affected by reasonable changes in prior specification. Detailed results are provided in Table 3.

Table 3 Prior sensitivity analysis comparing baseline and alternative prior specifications

| Prior specification | Spearman correlation with baseline | Top 10 hotspot overlap with baseline |
| --- | --- | --- |
| Baseline prior | 1 | 10 |
| Alternative prior for $\alpha_{NB}$ | 0.9996861991222057 | 10 |
| Alternative prior for $\sigma_{s}$ | 0.9996565152553872 | 10 |

5. **Prior Sensitivity Analysis**

#### 5.1. Scope of the sensitivity analysis

We conducted a systematic prior sensitivity analysis covering all seven prior distributions in the model as well as one distributional assumption, yielding eight alternative specifications in addition to the baseline model (nine full MCMC runs in total). The parameters examined were the intercept, covariate regression coefficients, the scale of the shared spatial effect, the shared loading scale, the scale of the disease-specific spatial effect, the distributional form of the disease-specific spatial effect, the innovation variance of the temporal RW1 process, and the negative binomial overdispersion parameter.

A one-at-a-time (OAT) strategy was adopted, such that only one prior was modified in each sensitivity run while all other specifications were kept at their baseline settings. This design allowed posterior differences to be attributed to a single prior adjustment.

#### 5.2. Alternative prior specifications

For each parameter, we selected an alternative specification that was more diffuse than the baseline prior. The eight alternative runs were as follows: alpha_d_norm05, gamma_norm02, sigma_s_hn02, lambda_d_hn02, sigma_sd_hn02, spatial_disease_t5, sigma_rw_hn10, and alpha_nb_g101.

The corresponding baseline and alternative specifications were(see Table 4 for more details):

Table 4 Baseline and alternative prior specifications for the prior sensitivity analysis

| **Parameter** | **Baseline prior** | **Alternative prior** |
| --- | --- | --- |
| alpha_d | Normal(0, 2) | Normal(0, 5) |
| gamma | Normal(0, 1) | Normal(0, 2) |
| sigma_s | HalfNormal(1) | HalfNormal(2) |
| lambda_d | HalfNormal(1) | HalfNormal(2) |
| sigma_sd | HalfNormal(1) | HalfNormal(2) |
| spatial_disease | Normal(0, sigma_sd) | StudentT(ν = 5, 0, sigma_sd) |
| sigma_rw | HalfNormal(0.5) | HalfNormal(1) |
| alpha_nb | Gamma(2, 0.5) | Gamma(1, 0.1) |

Each MCMC run used 16 chains, 2,000 warm-up iterations, and 2,000 posterior draws per chain, with target_accept = 0.95, yielding 32,000 posterior samples per run. The analysis was based on 20,808 country-year-disease observations (204 countries × 34 years × 3 diseases), with six covariates. The ICAR neighborhood graph contained 493 adjacency pairs, with a scaling factor of 0.729.

#### 5.3. Sensitivity metrics

We quantified the deviation of each alternative run from the baseline using six robustness metrics:

Spearman’s rho for country-level posterior mean shared hotspot scores

Top-10 overlap in hotspot countries

Top-10 Jaccard index

Spearman’s rho for country-level posterior exceedance probabilities, P(spatial_shared > 0)

Mean absolute difference in hotspot scores

Mean absolute difference in exceedance probabilities

Table 5 Results of the prior sensitivity analysis

| **Run** | **Spearman rho (hotspot)** | **Top-10 overlap** | **Jaccard** | **Spearman rho (exceedance)** | **Mean abs diff (hotspot)** | **Mean abs diff (exceedance)** |
| --- | --- | --- | --- | --- | --- | --- |
| base | 1.00000 | 10/10 | 1.0000 | 1.00000 | 0.00000 | 0.00000 |
| alpha_d_norm05 | 0.99969 | 10/10 | 1.0000 | 0.99961 | 0.00603 | 0.00366 |
| gamma_norm02 | 0.99975 | 10/10 | 1.0000 | 0.99953 | 0.00692 | 0.00382 |
| sigma_s_hn02 | 0.99966 | 10/10 | 1.0000 | 0.99938 | 0.16467 | 0.00472 |
| lambda_d_hn02 | 0.99957 | 10/10 | 1.0000 | 0.99942 | 0.19928 | 0.00531 |
| sigma_sd_hn02 | 0.99974 | 10/10 | 1.0000 | 0.99952 | 0.01002 | 0.00413 |
| spatial_disease_t5 | 0.97959 | 8/10 | 0.6667 | 0.96130 | 0.25290 | 0.07912 |
| sigma_rw_hn10 | 0.99972 | 10/10 | 1.0000 | 0.99938 | 0.00682 | 0.00376 |
| alpha_nb_g101 | 0.99969 | 10/10 | 1.0000 | 0.99951 | 0.00689 | 0.00407 |

#### 5.4. Key findings

Overall robustness was high. Across the eight alternative specifications, all Spearman rank correlations were greater than 0.96, and for seven of the eight alternatives the hotspot and exceedance rankings were nearly identical to the baseline (rho generally > 0.9995). The Top-10 hotspot list was unchanged in seven of the eight runs.

The core hotspot country set was fully preserved in those seven runs: ERI (Eritrea), MHL (Marshall Islands), BDI (Burundi), TWN (Taiwan), BGD (Bangladesh), TUV (Tuvalu), ZAF (South Africa), RWA (Rwanda), NRU (Nauru), and SWZ (Eswatini). Only their relative ordering changed slightly.

For sigma_s_hn02 and lambda_d_hn02, the mean absolute hotspot difference was comparatively larger (0.16467 and 0.19928, respectively), while the rank correlations remained extremely high and the Top-10 lists were unchanged. This indicates that these prior changes mainly affected the absolute scale of the hotspot scores rather than materially changing the relative ranking of countries.

The most noticeable deviation from the baseline was observed under spatial_disease_t5, in which the disease-specific spatial effect was assigned a heavier-tailed Student-t prior. In this run, TWN (Taiwan) and RWA (Rwanda) dropped out of the baseline Top-10 list, whereas LSO (Lesotho) and PRK (Democratic People’s Republic of Korea) entered it. This suggests that under a heavier-tailed prior for the disease-specific spatial effect, some movement may occur among the highest-ranked countries, although the overall ranking structure and broad spatial pattern remain largely preserved.

#### 5.5. Conclusion

Across the eight alternative prior specifications, the country-level hotspot rankings remained highly consistent with the baseline model, with Spearman correlations above 0.96 and a stable set of major hotspot countries in most runs. These findings indicate that the substantive conclusions were broadly robust to the tested prior specifications. However, some sensitivity was observed under the heavier-tailed Student-t prior for the disease-specific spatial effect, mainly in the composition of the highest-ranked hotspot countries. In addition, all sensitivity runs showed imperfect MCMC diagnostics, including maximum tree depth warnings and R-hat/effective sample size warnings for some parameters; therefore, these robustness results should be interpreted with appropriate caution.
